# Reproductive History and Age at Menarche in Relation to Reproductive Lifespan and Natural Menopause: A population-based analysis from NHANES 1999–March 2020

**DOI:** 10.1101/2025.11.13.25340186

**Authors:** Jingjing Feng, Jiayan Zhang, Jiahao Cai, Gaopi Deng

## Abstract

**Background:** To evaluate the associations of reproductive history(gravidity, parity, pregnancy loss) and age at menarche (AAM) with age at natural menopause (ANM) and reproductive lifespan (RL = ANM − AAM), and to assess effect modification and nonlinearity using a design-based, multiple-imputation framework.

**Methods:** We analyzed data from in NHANES 1999–March 2020 using stratified, clustered, survey-weighted methods with five imputations pooled by Rubin’s rules. We included women aged ≥60 years with self-reported AAM, ANM, gravidity and parity, excluding surgical or induced menopause. Primary outcomes were RL and ANM. Main analyses used linear models under two strategies: independent effects and mutual adjustment. Nonlinearity was assessed with restricted cubic splines (RCS) via joint Wald tests and AIC; pregnancy loss (approximately 65% zeros) was additionally evaluated using two-part models. Prespecified effect modifiers were income, race/ethnicity, and smoking; exploratory modifiers underwent BH-FDR control.

**Results:** The analytic sample included 3,167 participants. AAM showed a robust linear inverse association with RL (fully adjusted β = −0.871 years per later menarche year; 95% CI −1.011 to − 0.731; FDR p<0.001) and a borderline positive association with ANM (β = 0.129; 95% CI −0.011 to 0.269; p=0.07). Overall, parity and pregnancy loss were not significantly associated with RL or ANM across adjustment tiers. Two-part models indicated no transition effect and no dose–response among pregnancy-loss >0. Smoking modified the parity→ RL association (prespecified joint Wald p=0.026). Income displayed a nonlinear association with RL, with larger gains from low to middle income.

**Discussion:** In nationally representative data using rigorous design-based and multiple-imputation analyses, AAM was a stable determinant of RL with an approximately linear inverse association. In contrast, independent effects of gravidity, parity, and pregnancy loss on ANM/RL were limited. Lifestyle factors(smoking) and socioeconomic context modified specific associations. Confirmation in longitudinal cohorts with biomarker endpoints is needed to establish causality.

## Introduction

Natural menopause is defined as 12 consecutive months without menses, marking the age at natural menopause (ANM) and the end of the reproductive lifespan (RL). Age at menarche (AAM) and ANM are core indicators of female reproductive history, but they vary widely across global geographic, cultural, and socioeconomic settings.[1, 2] ANM is influenced by socioeconomic, lifestyle, and genetic factors, including family history of early menopause, education, occupation, smoking and alcohol use.[2–4] Epidemiologic studies indicate that later ANM and longer RL are associated with higher survival and longevity, each 1-year delay in ANM corresponds to about a 2% lower all-cause mortality risk. Conversely, earlier menopause is associated with increased risks of cardiovascular disease[5, 6], poorer cognition, depression[7], accelerated biological aging[8], increasing BMI[9], loss of muscle mass[10], dementia[11], diabetes[12], respiratory disease and mortality[13], and metabolic syndrome.[14]

Conceptually, a woman’s reproductive life can be described as biological reproductive years (AAM to ANM), effective reproductive years (time with fecundity and sexual activity), and social reproductive exposure (time exposed to sexual activity). [15] Most studies focus on biological reproductive years, defined as RL by AAM and ANM. Previous findings on the relationships among AAM, ANM, and RL are inconsistent. Some studies suggest earlier menarche is associated with higher risk of earlier menopause,[16] while others consider AAM a strong marker of reproductive duration.[17] Biologically and demographically, RL defines the window for natural fecundity: fertility declines rapidly after age 37, and natural conception is rare after 45 years[18]. After menopause, sustained estrogen deficiency increases risks of neurocognitive, cardiovascular, and skeletal disorders.[19–21] Conversely, longer RL is associated with greater longevity.[22] Thus, RL serves not only as a timeframe for female reproductive capacity but also as a significant indicator of women’s health and lifespan.

Menarche, menopause, and pregnancy within RL are highly dependent on hormonal regulation, ovarian function, and the hypothalamic-pituitary-ovarian (HPO) axis. Within this framework, reproductive behaviors manifest as outcomes such as gravidity, parity, and pregnancy loss. These may alter ANM and RL by affecting ovulation patterns, follicular depletion rates, and endocrine homeostasis[23].

Over time, female ANM and RL show increasing trends, while AAM shows a earlier trend. In the US population, from 1959 to 2019, mean ANM increased from 48.4 to 49.9 years, RL extended from 35.0 to 37.1 years, and AAM decreased from 13.5 to 12.7 years,[24] aligning with current estimates of ANM around 49–50 years[25, 26].

Regarding the association between reproductive behavior and menopause, some studies suggest that higher parity may delay ANM and extend RL through ovulation suppression during pregnancy and slowed follicular depletion.[16, 27–29] However, some research indicates that the physiological costs of high gravidity and parity might accelerate ovarian aging or shorten post-reproductive lifespan, leading to earlier ANM or shorter RL.[30] These effects may differ by socioeconomic and lifestyle characteristics. In addition, pregnancy loss may interact with early-life hormonal fluctuations, health stressors, and socioeconomic disadvantage such as education or access to care, potentially accelerating menopausal timing and shortening RL. At the molecular level, telomere length is linked to reproductive aging in women. Meanwhile, negative associations have been reported between telomeres and gravidity/parity[31–33], and shorter telomeres have been observed in women with recurrent pregnancy loss,[34] suggesting potential biological pathways linking “ reproductive burden” to reproductive aging.

Despite these insights, prior work on AAM–ANM–RL relations is inconsistent, and effects of reproductive history (gravidity, parity, pregnancy loss) may be confounded by socioeconomic and lifestyle factors[16, 27]. Nationally representative evidence using design-consistent methods is needed to clarify these relations and to evaluate effect modification and nonlinearity. We used NHANES 1999–March 2020 to examine associations of reproductive history and AAM with ANM and RL, and to assess modification by income, race/ethnicity, and smoking.

## Materials and methods

### Data source and study population

We used NHANES 1999–March 2020 (10 cycles), a multistage, stratified, clustered survey by the National Center for Health Statistics. We included women aged ≥60 years with self-reported AAM, ANM, gravidity, and parity, excluding surgical or induced menopause (hysterectomy, oophorectomy or exogenous estrogen use). The age ≥60 cutoff was applied to reduce survivor bias and late-recall biases for ANM. The publicly available datasets were accessed for research purposes in June 2025. The NHANES data are publicly available and de-identified; authors did not have access to information that could identify individual participants during or after data collection.

**Fig 1.**
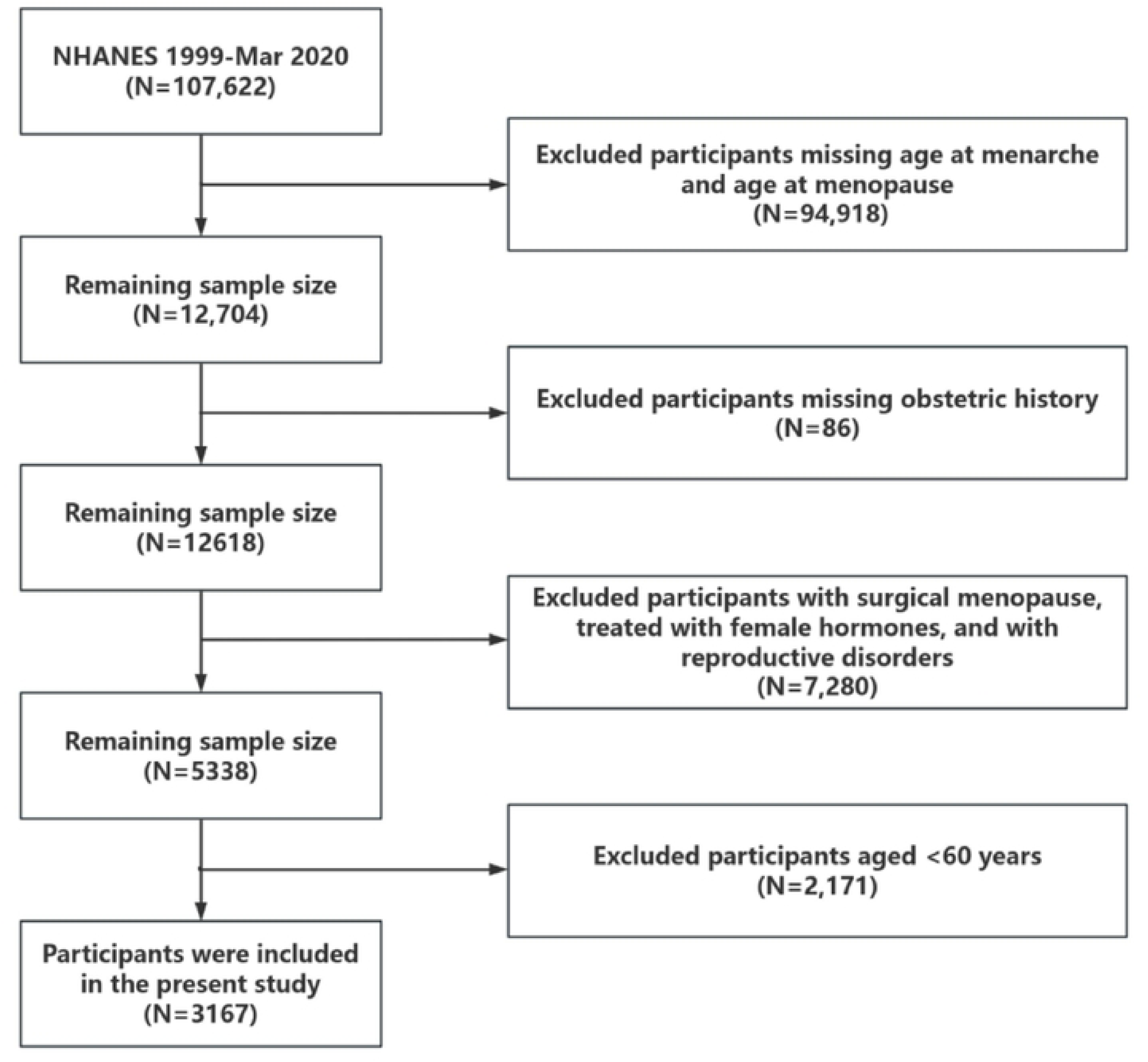
Flowchart of the participant selection from NHANES 1999-Mar 2020.

### Measures

Exposures included gravidity (lifetime number of pregnancies), parity (sum of vaginal and cesarean live births), pregnancy loss (calculated as gravidity − parity, encompassing miscarriages, stillbirths, ectopic or tubal pregnancies, and induced abortions), and AAM (age at first menstrual period). Primary outcomes were RL (RL = ANM − AAM) and ANM. To avoid structural coupling, AAM was not included as a covariate in RL models when assessing AAM effects[35].

### Covariates

Prespecified potential confounders included age, race/ethnicity (Mexican American, Other Hispanic, Non-Hispanic White, Non-Hispanic Black, Other Race), education level (Less than High school, High school or General Educational Development [15], Above High school), poverty-to-income ratio (PIR), marital status (Married or with partner vs. single), BMI (categorized as normal weight <25 kg/m², overweight 25–29.9 kg/m², obese ≥30 kg/m²), binge drinking status, smoking status (never, former, current), hypertension, diabetes, and cancer. Variable definitions followed NHANES documentation. PIR was also analyzed for potential nonlinearity as a secondary exposure.

### Statistical Analysis

All analyses followed NHANES analytical guidelines with strata, clusters, and MEC weights (combined across cycles).[36–38] Missing data were handled by multiple imputation (m=5 imputations) and pooled via Rubin’s rules[39]. Two-sided α=0.05, exploratory multiplicity was controlled by the Benjamini–Hochberg false discovery rate (FDR).[40] All statistical analyses were conducted in R version 4.3.0 using the survey package (version 4.2-1) for complex survey design analysis.

### Baseline comparisons

Participants were categorized by Parity (0, 1–2, ≥3). Categorical variables were compared using survey-weighted chi-square tests (F-adjusted), reported as counts and weighted percentages. Continuous variables, confirmed non-normal by the Shapiro–Wilk test, were compared using the weighted Kruskal–Wallis test (based on rank regression).

### Basic Regression Models

Two modeling strategies were employed. Strategy A (independent effects): each exposure (parity, gravidity, pregnancy loss, age at menarche) was modeled separately, with covariates added stepwise from unadjusted to fully adjusted models (age, race/ethnicity, education, income, marital status, BMI, alcohol, smoking, hypertension, diabetes, cancer). Strategy B (joint control): multiple exposures were included in the same model. Results were reported as β coefficients, standard errors (SE), 95% confidence intervals (CI), and p-values for each covariate adjustment level.

### Interaction analysis

Within survey-weighted linear regression models, product terms (X×G) were created for core exposures (X: Age at menarche, Parity, Pregnancy loss) and grouping variables (G: Age >70 vs. 60–70; Income: PIR≤1 vs. 1–4 vs.≥4; Education: ≤High school vs. >High school). Models with and without the interaction term were compared using likelihood ratio tests. Reference groups were Age (>70 years), Income (PIR≤1), Education (≤High school). Interaction coefficients, 95% CIs, and p values were reported.

### Nonlinearity

For each exposure–outcome pair (Y∼X), survey-weighted linear models were fitted under three covariate sets: unadjusted; basic (education, race, BMI); and full (age, race, education, marital status, BMI, alcohol, smoking, hypertension, diabetes, cancer, PIR). To assess nonlinearity, restricted cubic splines (RCS) were used in the survey-weighted linear models. Knots were placed at weighted quantiles, and degrees of freedom (df) were selected using Akaike Information Criterion (AIC), and overall nonlinearity was evaluated using a joint Wald test of the spline terms.[41, 42] FDR was controlled by Benjamini–Hochberg. Non-linearity was considered present if p<0.05 and ΔAIC ≤−2. We reported β, 95% CI, p values, and ΔAIC (linear vs RCS). Sensitivity analyses included alternative knot locations and df, weight trimming (0–5%), GAM validation of curve shapes (k=4,bs=’cr’)[43], and racial/ethnic stratification. Results were considered robust if effect direction and curve shape remained consistent across specifications. Because pregnancy loss had a high proportion of zero values (approximately 65%), a two-part approach was used. (i) transition effect (PL>0 vs 0) and (ii) dose–response within PL>0 using linear or RCS models, reverting to linear if RCS was unstable.

### Subgroup and sensitivity analyses

Outcomes were RL and ANM. The main exposure was parity (continuous), also categorized as 0, 1–2, and ≥3 with ordinal trend tests. Main model covariates included race/ethnicity, education (<High school/High school or equivalent/>High school), marital status (married/cohabiting vs single), smoking (never/former/current), continuous poverty income ratio (PIR), and survey cycle fixed effects (factor (SDDSRVYR)). The ANM model additionally adjusted for age at menarche. Survey-weighted generalized linear models were fitted in each imputed dataset, and estimates were combined with Rubin’s rules, and β, 95% CIs, and p values were reported. Joint Wald tests were used for interactions and spline non-linearity. Pre-specified effect Modifiers were Income (PIR ≤1, 1–4, ≥4), race/ethnicity, and smoking status. Joint Wald interaction tests for Parity were performed within both the Reproductive Lifespan and ANM models. Stratified forest plots were presented for significant interactions or those of public health relevance. Three-knot RCS were fitted for PIR→ RL and parity→ RL, we reported non-linearity p values and predicted contrasts at key percentiles/anchors (P10/P50/P90 and 0.5/1.5/4.0). Exploratory interactions for non-pre-specified variables (education, marital status, BMI, age dichotomy) were analyzed with outcome-specific FDR control and results are presented in the appendix. Additional sensitivity analyses using gravidity and pregnancy loss as alternative exposures; adjusting for potential mediators (BMI, binge drinking, hypertension, diabetes, cancer); replacing survey cycle fixed effects with birth cohort; restricting age ≤ 75 years to reduce survivor bias; reporting design degrees of freedom (df), weighted population size, and Kish’s effective sample size.

## Results

### Baseline characteristics

The analytic sample included 3,167 women aged ≥60 years. Table 1 presents baseline characteristics stratified by parity group (nulliparous [0], low parity [1–2], high parity [≥3]). Across parity groups (0; 1–2; ≥3), gravidity rose as expected with higher parity (mean ± SE: 0.32 ± 0.04; 2.29 ± 0.05; 5.15 ± 0.09; p<0.001), and pregnancy loss was modestly higher in the high-parity group (0.32 ± 0.04; 0.58 ± 0.04; 0.62 ± 0.04; p=0.003). High-parity women were older (RIDAGEYR: 69.07 ± 0.53; 68.97 ± 0.34; 71.05 ± 0.24; p<0.001), reported later age at menarche (AAM: 12.71 ± 0.12; 12.97 ± 0.06; 13.07 ± 0.05; p=0.010), and had lower income-to-poverty ratio (PIR: 2.69 ± 0.10; 2.88 ± 0.08; 2.32 ± 0.06; p<0.001).

Marked gradients by race/ethnicity and education were observed (both p<0.001), with non-Hispanic White and higher-educated women more often nulliparous or low parity and non-Hispanic Black/Hispanic and lower-educated women more often high parity. Diabetes prevalence differed across parity (p=0.001), whereas smoking, hypertension, cancer, and BMI did not significantly differ. Binge drinking differences were borderline (p=0.051). Neither reproductive lifespan (RL) nor age at natural menopause (ANM) differed significantly by parity group (RL p=0.227; ANM p=0.174), suggesting that crude associations may be confounded by sociodemographic and lifestyle factors.

**Table 1.** The baseline characteristics according to Parity Group.

| Variable | Nulliparous (0) | Low Parity (1-2) | High Parity ($\geq 3$) | Total | P-value |
| --- | --- | --- | --- | --- | --- |
| Gravidity | $0.32 \pm 0.04$ | $2.29 \pm 0.05$ | $5.15 \pm 0.09$ | $3.51 \pm 0.07$ | $< 0.001$ |
| Parity | $0.00 \pm 0.00$ | $1.71 \pm 0.02$ | $4.53 \pm 0.08$ | $2.94 \pm 0.07$ | $< 0.001$ |
| Pregnancy loss | $0.32 \pm 0.04$ | $0.58 \pm 0.04$ | $0.62 \pm 0.04$ | $0.57 \pm 0.02$ | 0.003 |
| Reproductive lifespan (years) | $36.70 \pm 0.48$ | $37.00 \pm 0.26$ | $36.46 \pm 0.18$ | $36.69 \pm 0.14$ | 0.227 |
| Age of menarche | $12.71 \pm 0.12$ | $12.97 \pm 0.06$ | $13.07 \pm 0.05$ | $12.99 \pm 0.04$ | 0.010 |
| Age of menopause | $49.41 \pm 0.44$ | $49.97 \pm 0.25$ | $49.53 \pm 0.18$ | $49.68 \pm 0.14$ | 0.174 |
| Age (years) | $69.07 \pm 0.53$ | $68.97 \pm 0.34$ | $71.05 \pm 0.24$ | $70.05 \pm 0.21$ | $< 0.001$ |
| PIR (unitless) | $2.69 \pm 0.10$ | $2.88 \pm 0.08$ | $2.32 \pm 0.06$ | $2.57 \pm 0.05$ | $< 0.001$ |
| Race | | | | | $< 0.001$ |
| Mexican American | 42 (3.1%) | 84 (2.5%) | 438 (7.7%) | 564 (5.2%) |  |
| Other Hispanic | 38 (5.4%) | 114 (5.7%) | 212 (5.9%) | 364 (5.8%) |  |
| Non-Hispanic white | 165 (78.4%) | 463 (75.9%) | 665 (66.8%) | 1293 (71.6%) |  |
| Non-Hispanic Black | 64 (7.8%) | 205 (8.6%) | 387 (12.1%) | 656 (10.3%) |  |
| Other Race | 27 (5.4%) | 105 (7.3%) | 158 (7.5%) | 290 (7.1%) |  |
| Education level |  |  |  |  | <0.001 |
| Less than high school | 79 (14.0%) | 258 (18.9%) | 925 (35.1%) | 1262 (26.6%) |  |
| High school or GED | 82 (26.2%) | 251 (28.6%) | 439 (30.9%) | 772 (29.4%) |  |
| Above high school | 175 (59.8%) | 462 (52.6%) | 496 (34.1%) | 1133 (44.0%) |  |
| Marital status |  |  |  |  | <0.001 |
| Married or with partner | 96 (31.1%) | 418 (50.3%) | 790 (45.9%) | 1304 (45.7%) |  |
| Single | 240 (68.9%) | 553 (49.7%) | 1070 (54.1%) | 1863 (54.3%) |  |
| Drinking status |  |  |  |  | 0.051 |
| Yes | 18 (5.9%) | 34 (2.4%) | 75 (3.5%) | 127 (3.4%) |  |
| No | 318 (94.1%) | 937 (97.6%) | 1785 (96.5%) | 3040 (96.6%) |  |
| Smoking status |  |  |  |  | 0.124 |
| Never | 207 (55.5%) | 584 (57.0%) | 1221 (62.5%) | 2012 (59.7%) |  |
| Former | 90 (33.0%) | 276 (31.2%) | 437 (26.1%) | 803 (28.8%) |  |
| Current | 39 (11.5%) | 111 (11.8%) | 202 (11.4%) | 352 (11.5%) |  |
| hypertension |  |  |  |  | 0.121 |
| Yes | 172 (50.4%) | 589 (56.9%) | 1129 (58.9%) | 1890 (57.1%) |  |
| No | 164 (49.6%) | 382 (43.1%) | 731 (41.1%) | 1277 (42.9%) |  |
| diabetes |  |  |  |  | 0.001 |
| Yes | 50 (11.7%) | 182 (15.6%) | 495 (20.9%) | 727 (17.8%) |  |
| No | 286 (88.3%) | 789 (84.4%) | 1365 (79.1%) | 2440 (82.2%) |  |
| cancer |  |  |  |  | 0.885 |
| Yes | 50 (17.3%) | 144 (17.8%) | 232 (16.9%) | 426 (17.3%) |  |
| No | 286 (82.7%) | 827 (82.2%) | 1628 (83.1%) | 2741 (82.7%) |  |
| BMI |  |  |  |  | 0.838 |
| Normal weight (<25kg/m<sup>2</sup>) | 96 (28.3%) | 249 (26.3%) | 465 (27.4%) | 810 (27.1%) |  |
| Overweight (25–29.9kg/m<sup>2</sup>) | 112 (29.8%) | 311 (34.1%) | 577 (30.9%) | 1000 (31.9%) |  |
| Obese (≥30kg/m<sup>2</sup>) | 128 (41.9%) | 411 (39.6%) | 818 (41.8%) | 1357 (41.0%) |  |

Values are survey-weighted. Continuous variables are mean ± SE; categorical variables are n (weighted %). Tests: F-adjusted $\chi^2$ (categorical) and weighted rank-based tests (continuous). Abbreviations: AAM, age at menarche; ANM, age at natural menopause; RL, reproductive lifespan; PIR, poverty-to-income ratio; BMI, body mass index; SE, standard error.

### Main associations

Independent-effect models (Table 2) showed a robust inverse association of AAM with RL across all covariate tiers. Point estimates were highly consistent from unadjusted to fully adjusted specifications (e.g., RL per 1-year later menarche: β = −0.958; 95% CI −1.098, −0.818; to β = − 0.871; 95% CI −1.011, −0.731; FDR p<0.001). For ANM, AAM was borderline positive in the fully adjusted model (β = 0.129; 95% CI −0.011to 0.269; p=0.07) but non-significant at lower adjustment tiers.

Parity was inversely associated with RL before adjustment (β = −0.169; 95% CI −0.311 to − 0.028; p=0.019) but attenuated to null with additional covariates (Models 2–5; e.g., Model 5 β = 0.011; p=0.887). Pregnancy loss showed no significant association with RL or ANM at any tier. Mutually adjusted models (Table 3) yielded materially similar conclusions: AAM remained strongly and inversely related to RL (e.g., Model 5 β = −0.873; p<0.001), while parity and pregnancy loss showed no independent linear associations with either RL or ANM after adjustment for the other exposures and covariates.

**Table 2.** Survey-weighted linear regression (independent effects) of exposures on outcomes.

| Outcome | Predictor | Model 1 |  | Model 2 |  | Model 3 |  | Model 4 |  | Model 5 |  |
| --- | --- | --- | --- | --- | --- | --- | --- | --- | --- | --- | --- |
| | | $\beta$ (95%CI) | P | $\beta$ (95%CI) | P | $\beta$ (95%CI) | P | $\beta$ (95%CI) | P | $\beta$ (95%CI) | P |
| Age at menopause | Age of menarche | 0.042 |  | 0.066 |  | 0.096 |  | 0.122 |  | 0.129 |  |
|  |  | (-0.098, | 0.558 | (-0.078, | 0.368 | (-0.045, | 0.183 | (-0.018, | 0.089 | (-0.011, | 0.07 |
|  |  | 0.182) |  | 0.209) |  | 0.238) |  | 0.263) |  | 0.269) |  |
| | | $\beta$ (95%CI) | P | $\beta$ (95%CI) | P | $\beta$ (95%CI) | P | $\beta$ (95%CI) | P | $\beta$ (95%CI) | P |
| Age at menopause | Parity | -0.096 |  | -0.014 |  | 0.045 |  | 0.046 |  | 0.042 |  |
|  |  | (-0.234, 0.042) | 0.171 | (-0.160, 0.132) | 0.850 | (-0.107, 0.198) | 0.558 | (-0.106, 0.199) | 0.552 | (-0.110, 0.194) | 0.588 |
| Age at menopause | Pregnancy loss | 0.160 |  | 0.161 |  | 0.158 |  | 0.163 |  | 0.162 |  |
|  |  | (-0.108, 0.427) | 0.242 | (-0.106, 0.428) | 0.236 | (-0.095, 0.411) | 0.220 | (-0.091, 0.418) | 0.208 | (-0.092, 0.415) | 0.211 |
| Reproductive lifespan | Age of menarche | -0.958 |  | -0.934 |  | -0.904 |  | -0.878 |  | -0.871 |  |
|  |  | (-1.098, -0.818) | 0.001 | (-1.078, -0.791) | 0.001 | (-1.045, -0.762) | 0.001 | (-1.018, -0.737) | 0.001 | (-1.011, -0.731) | 0.001 |
| Reproductive lifespan | Parity | -0.169 |  | -0.067 |  | 0.018 |  | 0.018 |  | 0.011 |  |
|  |  | (-0.311, -0.028) | 0.019 | (-0.219, 0.085) | 0.387 | (-0.139, 0.175) | 0.822 | (-0.139, 0.176) | 0.818 | (-0.145, 0.167) | 0.887 |
| Reproductive lifespan | Pregnancy loss | 0.145 |  | 0.150 |  | 0.142 |  | 0.147 |  | 0.144 |  |
|  |  | (-0.117, 0.408) | 0.279 | (-0.112, 0.411) | 0.262 | (-0.104, 0.388) | 0.257 | (-0.098, 0.392) | 0.239 | (-0.099, 0.388) | 0.245 |

Survey-weighted, MI-pooled linear models (Gaussian) estimating independent associations of each exposure with each outcome. Models 1–5: Model 1, minimal adjustment; Model2, plus age, race; Model 3, plus sociodemographics (education, income-to-poverty ratio, marital status); Model 4, plus lifestyle factors (BMI, binge drinking, smoking status); Model 5, plus comorbidities (hypertension, diabetes, cancer). Results are $\beta$ , SE, 95% CI, p.

**Table 3.**
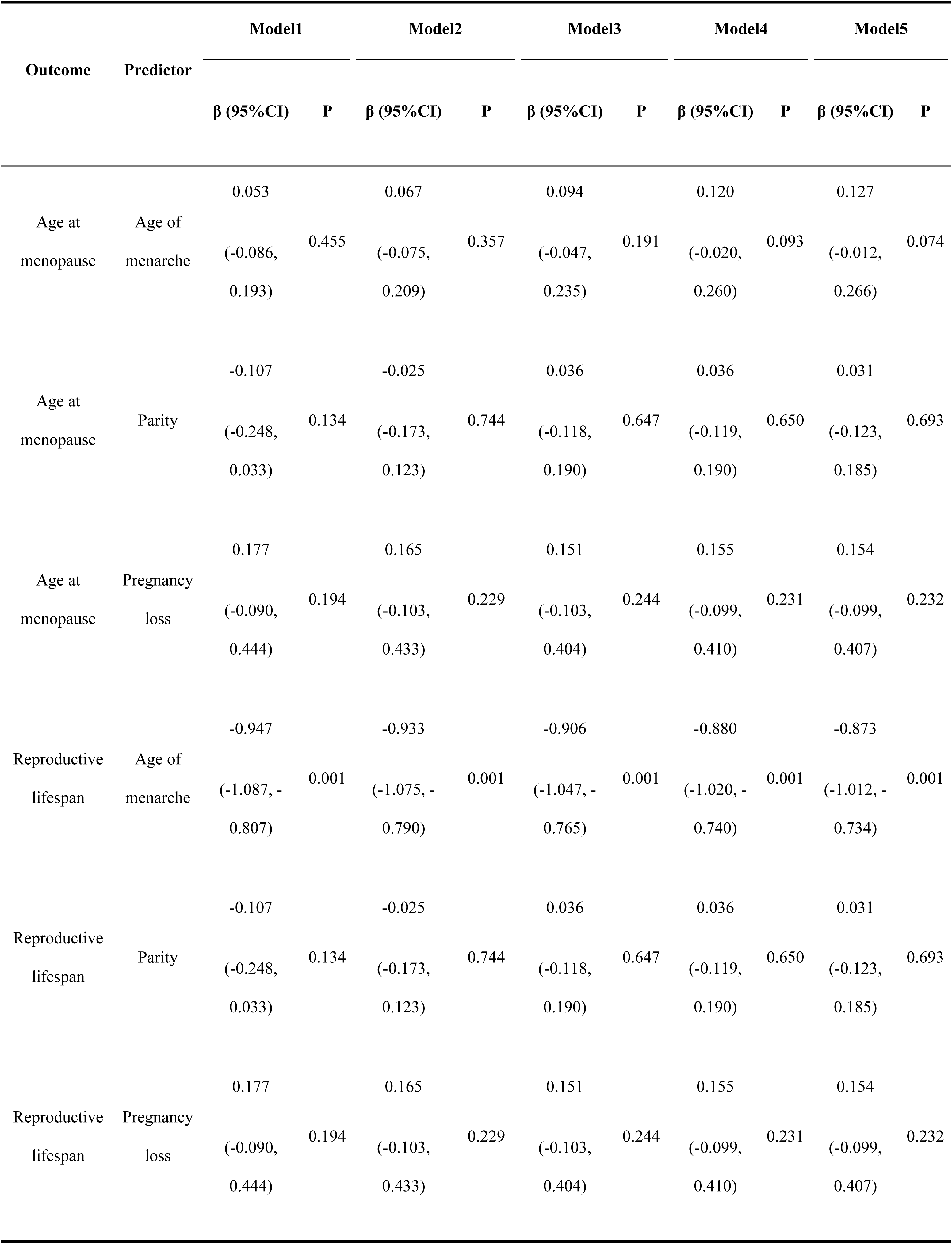
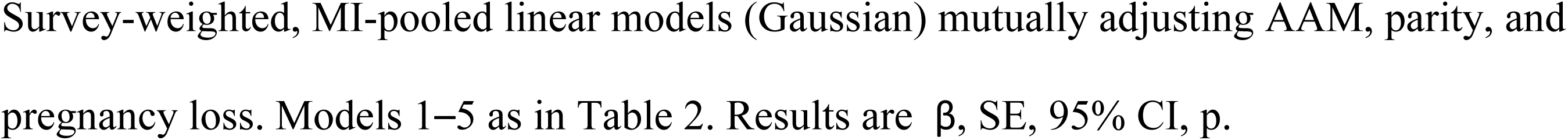
Survey-weighted linear regression (mutual adjustment) of exposures on outcomes.

| Outcome | Predictor | Model1 |  | Model2 |  | Model3 |  | Model4 |  | Model5 |  |
| --- | --- | --- | --- | --- | --- | --- | --- | --- | --- | --- | --- |
| | | $\beta$ (95%CI) | P | $\beta$ (95%CI) | P | $\beta$ (95%CI) | P | $\beta$ (95%CI) | P | $\beta$ (95%CI) | P |
| Age at menopause | Age of menarche | 0.053 |  | 0.067 |  | 0.094 |  | 0.120 |  | 0.127 |  |
|  |  | (-0.086, | 0.455 | (-0.075, | 0.357 | (-0.047, | 0.191 | (-0.020, | 0.093 | (-0.012, | 0.074 |
|  |  | 0.193) |  | 0.209) |  | 0.235) |  | 0.260) |  | 0.266) |  |
| Age at menopause | Parity | -0.107 |  | -0.025 |  | 0.036 |  | 0.036 |  | 0.031 |  |
|  |  | (-0.248, | 0.134 | (-0.173, | 0.744 | (-0.118, | 0.647 | (-0.119, | 0.650 | (-0.123, | 0.693 |
|  |  | 0.033) |  | 0.123) |  | 0.190) |  | 0.190) |  | 0.185) |  |
| Age at menopause | Pregnancy loss | 0.177 |  | 0.165 |  | 0.151 |  | 0.155 |  | 0.154 |  |
|  |  | (-0.090, | 0.194 | (-0.103, | 0.229 | (-0.103, | 0.244 | (-0.099, | 0.231 | (-0.099, | 0.232 |
|  |  | 0.444) |  | 0.433) |  | 0.404) |  | 0.410) |  | 0.407) |  |
| Reproductive lifespan | Age of menarche | -0.947 |  | -0.933 |  | -0.906 |  | -0.880 |  | -0.873 |  |
|  |  | (-1.087, - | 0.001 | (-1.075, - | 0.001 | (-1.047, - | 0.001 | (-1.020, - | 0.001 | (-1.012, - | 0.001 |
|  |  | 0.807) |  | 0.790) |  | 0.765) |  | 0.740) |  | 0.734) |  |
| Reproductive lifespan | Parity | -0.107 |  | -0.025 |  | 0.036 |  | 0.036 |  | 0.031 |  |
|  |  | (-0.248, | 0.134 | (-0.173, | 0.744 | (-0.118, | 0.647 | (-0.119, | 0.650 | (-0.123, | 0.693 |
|  |  | 0.033) |  | 0.123) |  | 0.190) |  | 0.190) |  | 0.185) |  |
| Reproductive lifespan | Pregnancy loss | 0.177 |  | 0.165 |  | 0.151 |  | 0.155 |  | 0.154 |  |
|  |  | (-0.090, | 0.194 | (-0.103, | 0.229 | (-0.103, | 0.244 | (-0.099, | 0.231 | (-0.099, | 0.232 |
|  |  | 0.444) |  | 0.433) |  | 0.404) |  | 0.410) |  | 0.407) |  |

### Interaction Analysis

We evaluated 16 exposure-by-group interactions (age group, education, and income categories) (Table 4). Two nominal signals (p<0.05) appeared for pregnancy loss × high income (PIR ≥4) relative to low income (PIR ≤1): a more positive slope per additional loss for ANM (β=0.702; SE=0.306; p=0.022) and for RL (β=0.633; SE=0.322; p=0.050). However, neither survived BH-FDR correction, and no other prespecified interactions reached significance.

**Table 4.** Interaction analysis—Survey-weighted models with exposure-by-group terms.

| Outcome | Interaction | Beta | SE | P |
| --- | --- | --- | --- | --- |
| Age at menopause | Age of menarche x Age Group: middle-aged (60–70 years) | 0.094 | 0.149 | 0.526 |
| Age at menopause | Age of menarche x Education: high school or higher | 0.014 | 0.165 | 0.931 |
| Age at menopause | Parity x Age Group: middle-aged (60–70 years) | -0.137 | 0.109 | 0.208 |
| Age at menopause | Parity x Income: High income ($\geq 4$) | 0.108 | 0.182 | 0.553 |
| Age at menopause | Parity x Income: Middle income (1-4) | -0.040 | 0.140 | 0.777 |
| Age at menopause | Pregnancy loss x Age Group: middle-aged (60–70 years) | -0.220 | 0.233 | 0.344 |
| Age at menopause | Pregnancy loss x Income: High income ($\geq 4$) | 0.702 | 0.306 | 0.022 |
| Age at menopause | Pregnancy loss x Income: Middle income (1-4) | 0.033 | 0.214 | 0.878 |
| Reproductive lifespan | Age of menarche x Age Group: middle-aged (60–70 years) | 0.094 | 0.149 | 0.526 |
| Reproductive lifespan | Age of menarche x Education: high school or higher | 0.014 | 0.165 | 0.931 |
| Reproductive lifespan | Parity x Age Group: middle-aged (60–70 years) | -0.163 | 0.107 | 0.128 |
| Reproductive lifespan | Parity x Income: High income ($\geq 4$) | 0.116 | 0.189 | 0.540 |
| Reproductive lifespan | Parity x Income: Middle income (1-4) | 0.011 | 0.153 | 0.943 |
| Reproductive lifespan | Pregnancy loss x Age Group: middle-aged (60–70 years) | -0.276 | 0.246 | 0.264 |
| Reproductive lifespan | Pregnancy loss x Income: High income ($\geq 4$) | 0.633 | 0.322 | 0.050 |
| Reproductive lifespan | Pregnancy loss x Income: Middle income (1-4) | 0.138 | 0.244 | 0.571 |

Reference categories: age group: older ( $>70$ years); income group: low income ( $\leq 1$ ); education: below high school. Comparators: middle-aged (60–70 years), middle income (1–4), high income ( $\geq 4$ ). For interaction terms $X \times G$ , $\beta_3$ represents the difference in the $X \rightarrow Y$ slope for the comparator versus the reference group.

### Nonlinearity analysis

The AAM→RL association was consistently linear and significant across covariate sets (Table 5). Linear slopes ranged from β = −0.958 (95% CI −1.098 to −0.818) unadjusted to β = −0.871 (95% CI −1.011 to −0.731) fully adjusted (FDR p<0.001). Fig 2 visualizes the three linear fits (unadjusted/basic/full) with 95% CIs. Restricted cubic splines (RCS) suggested only mild curvature and did not improve model fit versus linear (ΔAIC= +1.4 to +1.6; joint Wald nonlinearity not met), with shape corroborated by generalized additive models (GAM) (Fig 3; Supplementary Tables S2–S5). Because about 65% of pregnancy-loss values were zero, we used a two-part strategy. After full adjustment, transition effects (loss>0 vs 0) were non-significant for both RL (β=0.37 years; SE=0.40; p=0.36) and ANM (β=0.47 years; SE=0.38; p=0.22); among loss>0, dose–response slopes were near zero and spline terms were non-significant, yielding approximately flat curves (Supplementary Fig S1). Comprehensive linear results for all 18 exposure–outcome combinations across three covariate sets (3 exposures × 2 outcomes × 3 sets) are reported in Supplementary Table S1. Supplementary Table S6 shows consistent directions by race/ethnicity, supporting robustness. Exploratory income interactions within the two-part framework were non-significant after FDR control (Supplementary Table S7).

**Fig 2.**
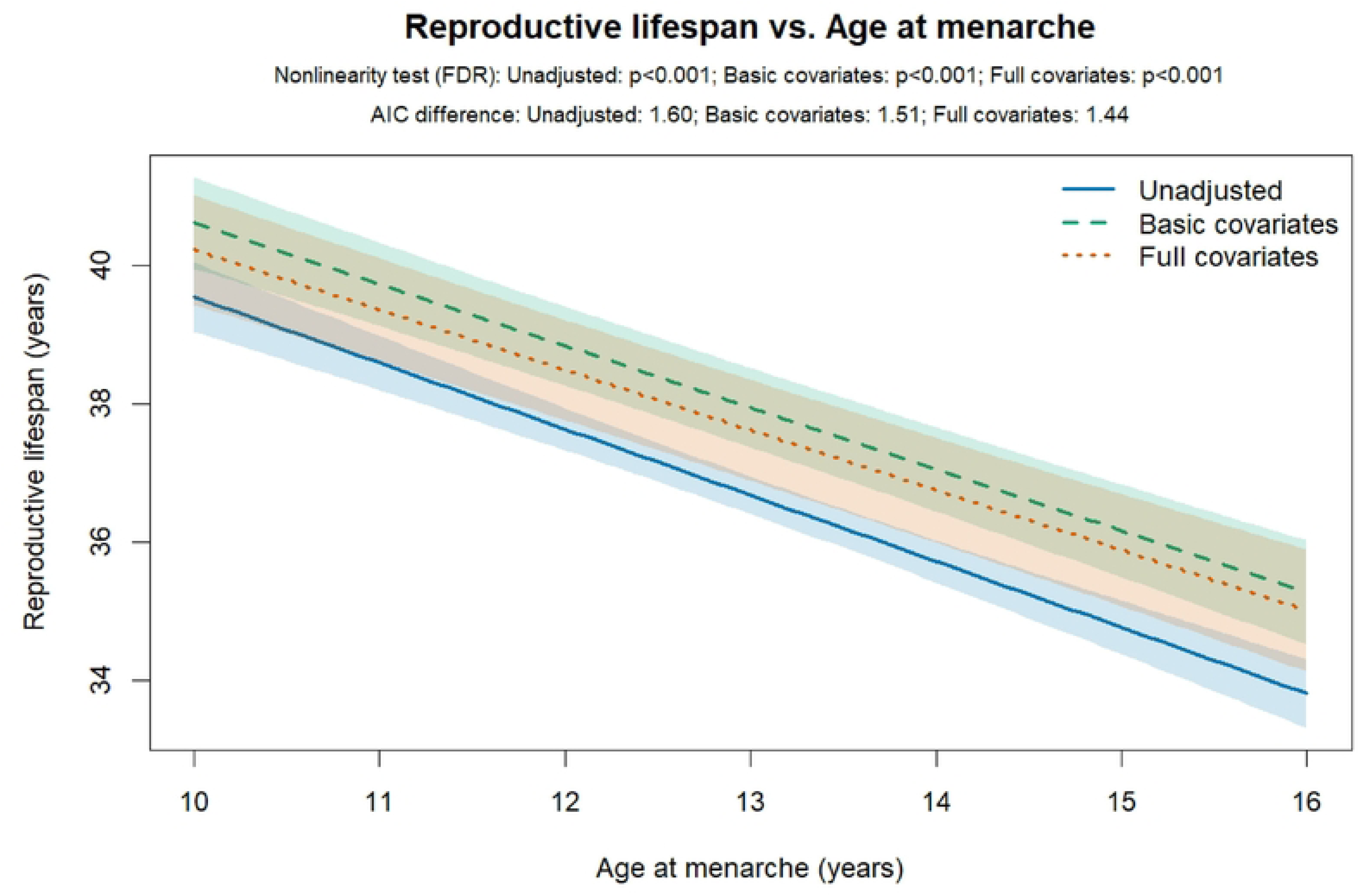
Linear relationship between Age of menarche and Reproductive lifespan under three covariate sets. Survey-weighted linear models pooled across five imputations. Lines represent unadjusted (solid), basic covariates (dashed), and fully adjusted (dotted); shaded bands are 95% CIs.

**Fig 3.**
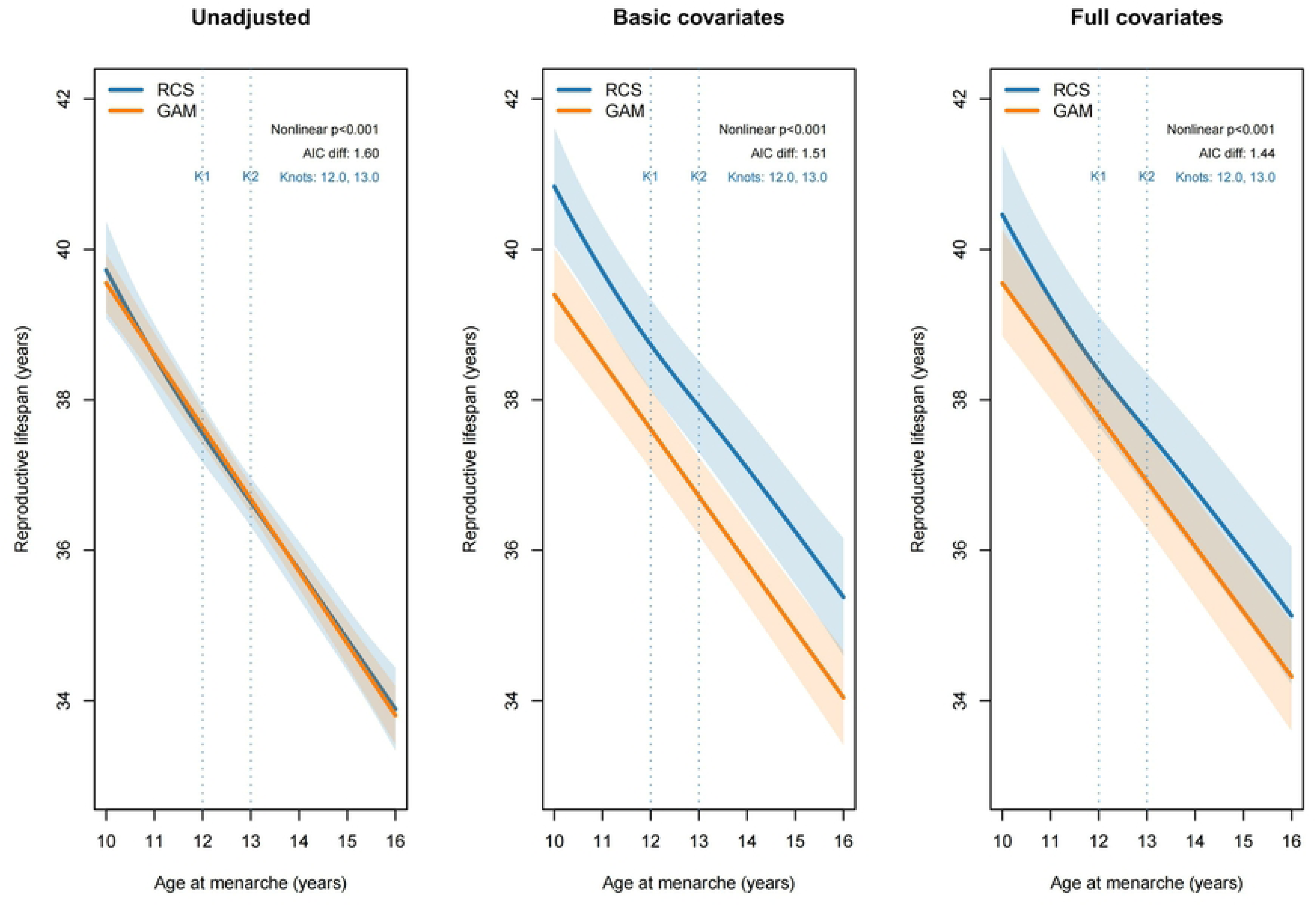
Nonlinearity diagnostics for Age of menarche vs. Reproductive lifespan: restricted cubic spline (RCS) versus generalized additive model (GAM). Three panels show unadjusted (A), basic covariates (B), and fully adjusted (C) models. Each panel overlays RCS and GAM fits (survey-weighted, MI-pooled) for comparison; bands are 95% CIs. Abbreviations: RCS, restricted cubic spline; GAM, generalized additive model.

**Table 5.** Linear association between Age of menarche and Reproductive lifespan across covariate sets.

| Exposure | Outcome | Covariateset | Beta(95%CI) | P | FDR |
| --- | --- | --- | --- | --- | --- |
| Age of menarche | Reproductive lifespan | Unadjusted | -0.958 (-1.098, -0.818) | <0.001 | <0.001 |
| Age of menarche | Reproductive lifespan | Basic | -0.893 (-1.032, -0.754) | <0.001 | <0.001 |
| Age of menarche | Reproductive lifespan | Fully adjusted | -0.871 (-1.011, -0.731) | <0.001 | <0.001 |

Covariate Sets: Unadjusted: No covariates included. Basic: Age, race/ethnicity, education, income-to-poverty ratio, marital status. Fully Adjusted: Basic covariates plus lifestyle and comorbidities: BMI, binge drinking, smoking status, hypertension, diabetes, cancer.

### Subgroup and Sensitivity Analyses

In our survey-weighted, multiply imputed linear models, parity showed no overall linear main effect on reproductive lifespan (RL: β=−0.002, p=0.976) or age at natural menopause (ANM: β =0.029, p=0.691). Ordinal trend tests across categories (0, 1-2, ≥3) was also non-significant (RL p=0.605; ANM p=0.434). Prespecified interactions indicated significant modification of the parity→ RL association by smoking (joint Wald p=0.026) and a borderline effect for ANM (p=0.077). The smoking-stratified forest plot (Fig 4) shows heterogeneous directions and uncertainty across strata, supporting the interaction. Supplementary Figs S2–S3 show no significant parity × income (three categories) or parity × race interactions. Restricted cubic splines (RCS) indicated significant nonlinearity for PIR→RL (Wald p=0.006; Fig 5), with larger gains from low to middle income: P50− P10=0.939 years (95% CI 0.245 to 1.633) versus P90−P50=0.654 years (95% CI −0.233 to 1.540).

Sensitivity analyses using gravidity or pregnancy loss instead of parity yielded generally consistent and non-significant results. Despite isolated stratum-specific associations suggesting heterogeneity (e.g., pregnancy loss with RL among former smokers), no interaction terms (education, marital status, BMI, age groups) were significant after FDR control. Design degrees of freedom (df) = 161; Kish effective sample size = 1,669; weighted population represented = 8.26 million (details in Supplementary Table S8).

**Fig 4.**
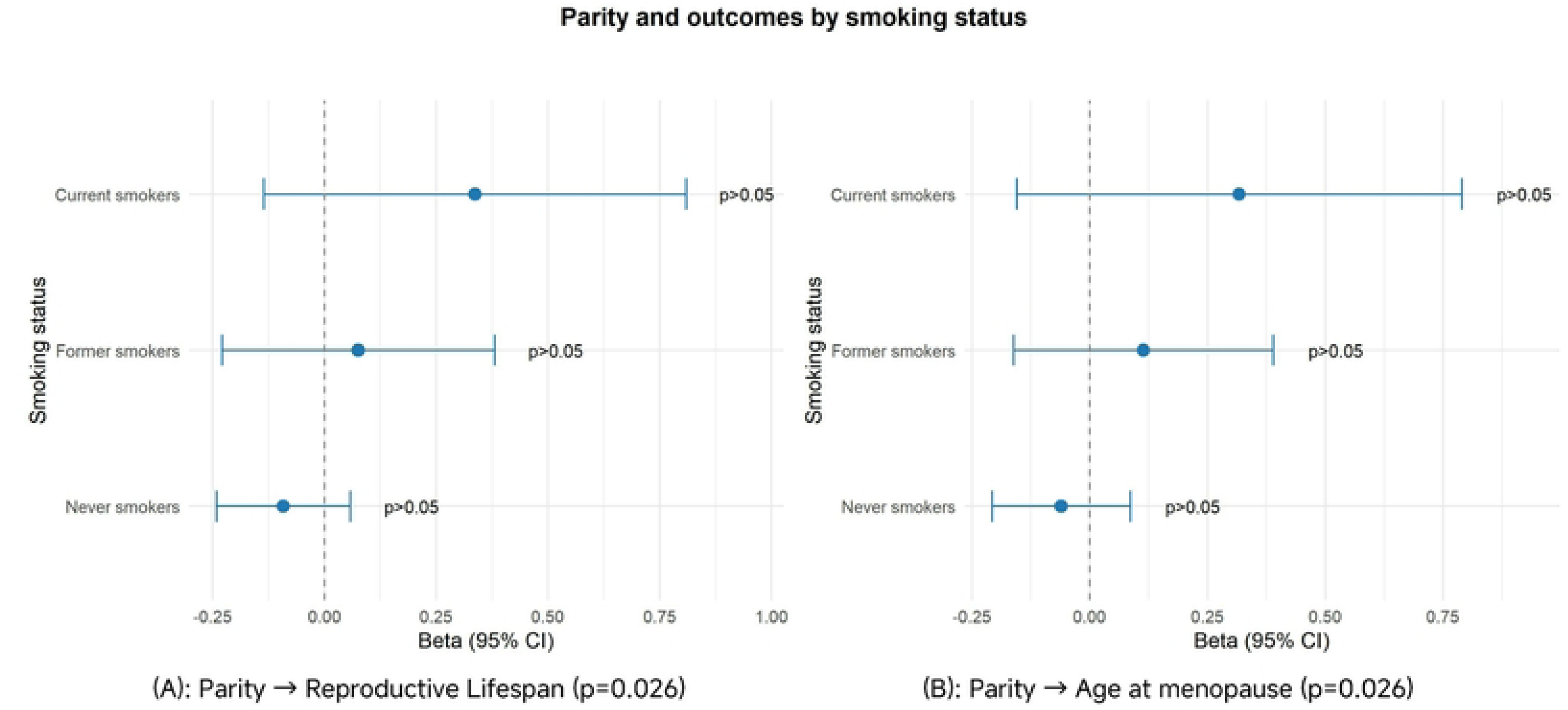
Parity and reproductive outcomes by smoking status. Two-panel forest plot of parity associations within smoking strata. Left: Reproductive Lifespan; Right: Age at menopause. Survey-weighted, MI-pooled models adjust for race, education, marriage, PIR, period fixed effects (Age at menopause models additionally adjust for age at menarche). Points show Beta and 95% CI.

**Fig 5.**
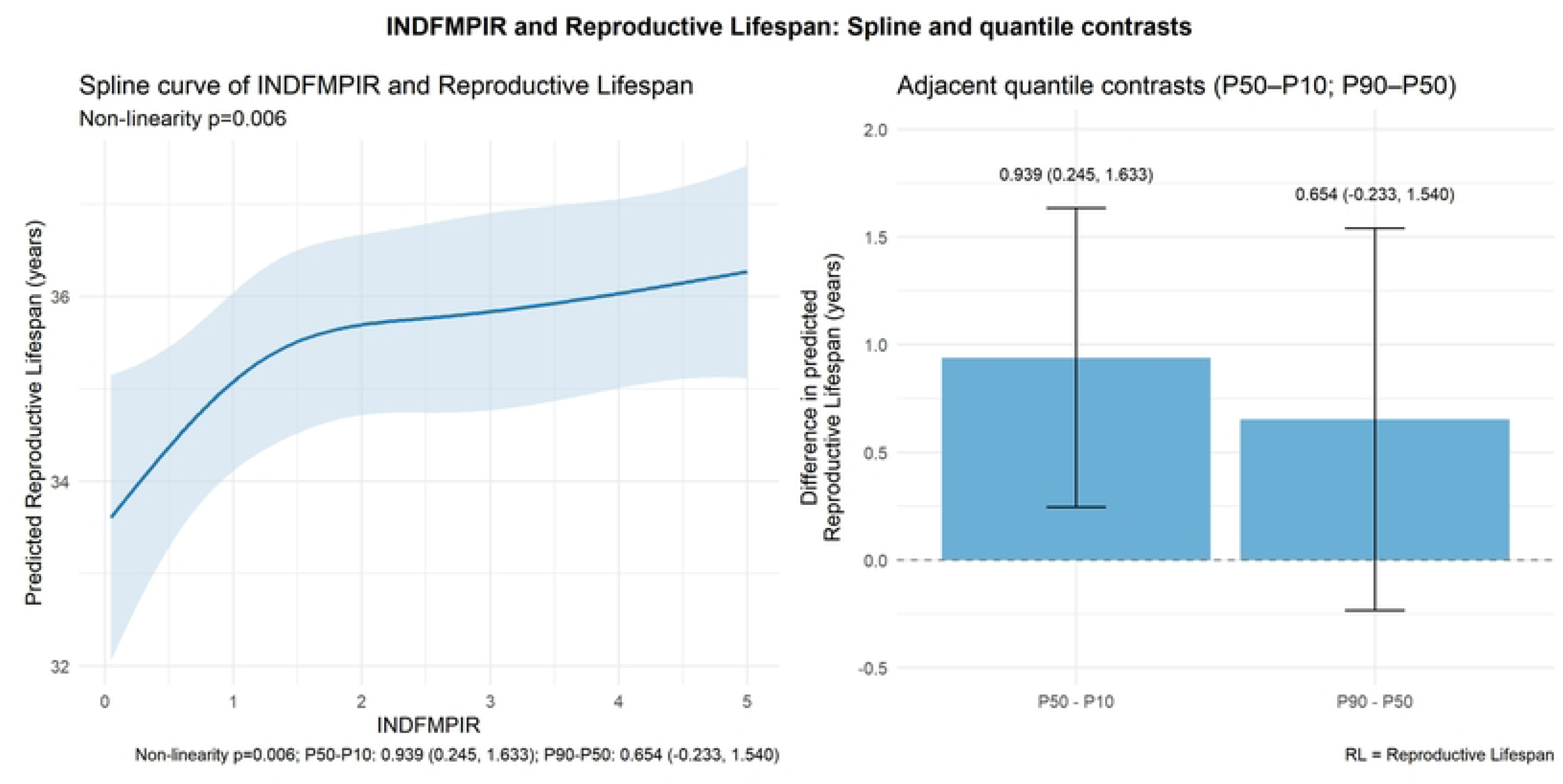
Restricted cubic spline of PIR with reproductive lifespan and adjacent quantile contrasts. Survey-weighted, MI-pooled models adjusted for prespecified covariates. nonlinearity p=0.006. Adjacent contrasts: P50−P10=0.939 (95% CI 0.245–1.633); P90−P50=0.654 (95% CI −0.233–1.540). RL = Reproductive Lifespan.

**Table 6.** Association of Parity with Reproductive Lifespan and Age at Menopause.

| Outcome | Exposure | Beta | SE | 95% CI | P value | Interaction p |  |  |
| --- | --- | --- | --- | --- | --- | --- | --- | --- |
|  |  |  |  |  |  | Income | Race | Smoking |
| Reproductive Lifespan | Parity (continuous) | -0.002 | 0.074 | -0.002 (-0.147, 0.143) | 0.976 | 0.906 | 0.401 | 0.026 |
| Reproductive Lifespan | Parity (trend) | 0.117 | 0.226 | 0.117 (-0.326, 0.560) | 0.605 | 0.405 | 0.2 | 0.277 |
| Age at menopause | Parity (continuous) | 0.029 | 0.073 | 0.029 (-0.114, 0.172) | 0.691 | 0.81 | 0.644 | 0.077 |
| Age at menopause | Parity (trend) | 0.171 | 0.218 | 0.171 (-0.257, 0.599) | 0.434 | 0.292 | 0.245 | 0.495 |

Survey-weighted, MI-pooled estimates. Models adjust for race/ethnicity, education, marital status, smoking, continuous PIR, and survey-cycle fixed effects; ANM models additionally adjust for AAM. Interaction p-values are joint Wald tests for Parity × Income (PIR $\leq 1$ , 1–4, $\geq 4$ ), Parity × Race, and Parity × Smoking. Results are $\beta$ , 95% CI, p. Abbreviations: RL, reproductive lifespan; ANM, age at natural menopause; PIR, poverty-to-income ratio.

## Discussion

This study used a nationally representative NHANES sample to assess the associations of reproductive history (gravidity, parity, pregnancy loss) and sexual maturation timing (age at menarche, AAM) with reproductive aging phenotypes (age at natural menopause, ANM; reproductive lifespan, RL). Within a complex survey framework with sampling weights and multiple imputation, we addressed zero inflation and nonlinearity and tested prespecified effect modification. Compared with studies that did not fully account for design effects and multiplicity, our approach provides greater methodological robustness and nationally representative estimates.

In our analyses, AAM showed a stable linear inverse association with RL (approximately −0.9 years per 1-year later menarche), consistent across two modeling strategies and all covariate sets and remaining significant after FDR control. AAM exhibited only borderline positively associated with ANM in fully adjusted models (β=0.13 years). In contrast, Parity and pregnancy loss showed no overall main effects on RL or ANM across adjustment tiers, including non-significant ordinal trends across parity categories (0, 1–2, ≥3). In two-part models for pregnancy loss, neither the transition effect (pregnancy loss >0 vs 0) nor the dose–response slope within pregnancy loss >0 was significant after full adjustment, and curves were approximately horizontal, suggesting limited average associations with RL/ANM after controlling for confounding or possible dilution by heterogeneity.

Overall, nonlinearity and sensitivity analyses supported the linear specification for AAM→RL. Although joint RCS tests suggested mild curvature, ΔAIC versus linear was positive (+1.4–1.6), failing the prespecified AND criterion (nonlinearity p<0.05 and ΔAIC≤−2). Thus, linear models were retained for primary inference and RCS presented as sensitivity. Alternative knot/df choices and weight trimming had minimal impact; GAM and RCS curves (with 95% CIs) largely overlapped; race/ethnicity strata were directionally consistent, supporting robustness. In contrast, AAM→ANM and gravidity/parity with either outcome were not significant after FDR control, full results are provided in Supplementary Table S1.

Regarding effect modification and social determinants, prespecified joint Wald tests indicated significant modification of the parity→ RL association by smoking status (interaction p=0.026) and a borderline effect for ANM (p=0.077). The smoking-stratified forest plot showed heterogeneous directions and uncertainty across strata, consistent with interaction. RCS indicated significant nonlinearity for PIR→RL (Wald p=0.006), with larger gains from low to middle income (P50− P10=0.94 years, 95% CI 0.25 to 1.63) than from middle to high income (P90−P50=0.65 years, 95% CI −0.23 to 1.54), suggesting that integrated interventions (e.g., nutrition, primary care access, and chronic disease management) may yield greater marginal benefits as resources increase from scarcity to adequacy. The exploratory income × pregnancy loss interaction was nominally significant within the two-part framework but non-significant after FDR control and should be interpreted cautiously (Supplementary Table S7).

Our findings of a robust inverse AAM→ RL association and a borderline positive AAM→ ANM association are consistent with prior evidence. Our finding of non-significant associations between reproductive factors and ANM/RL after comprehensive adjustment for confounders contrasts with some previous reports of positive associations. This discrepancy likely reflects methodological and population differences. We controlled for strong confounders including smoking, socioeconomic status, education, BMI, and comorbidities. Prior studies shows that parity→ ANM positive effects often attenuate or become non-significant after adjusting for these factors, suggesting residual confounding in earlier estimates[44, 45]. In addition, cross-country and cohort differences in contraception, breastfeeding, smoking, and education can alter the direction and magnitude of parity → ANM associations.[24] Our use of complex survey design, multiple imputation, FDR control, and explicit handling of nonlinearity and zero inflation yields nationally representative, design-consistent estimates; studies lacking design-consistent inference and multiplicity control may be more susceptible to nominal significance. Baseline differences in age, AAM, socioeconomic status, and race/ethnicity across parity groups likely confounded the unadjusted association, explaining the disappearance of nominal significance after full adjustment. [46]

Mechanistically, genetic and large population studies indicate ANM is primarily determined by intrinsic ovarian aging pathways (DNA damage response, homologous recombination repair, apoptosis, follicular depletion) and modifiable lifestyle factors (notably smoking), with limited net effects from pregnancy history[47]. U.S. secular evidence likewise links increases in ANM and RL and earlier AAM to cohort shifts in fertility, education, and smoking.[24] From a public health perspective, longer RL is associated with lower cardiovascular mortality.[48] Within this framework, our results support AAM as a more stable determinant of RL, whereas the independent effects of reproductive history appear modest at the population level and sensitive to confounding and population heterogeneity, consistent with synthesized evidence[2, 44, 45, 49].

We also observed significant modification of the parity→ RL association by smoking. Smoking has repeatedly been linked to earlier age at menopause through multiple pathways—pro-apoptotic and oxidative stress effects, aromatase inhibition and perturbed estrogen metabolism, anti-estrogenic actions, and toxicity to the follicular microenvironment—impairing ovarian reserve and the HPO axis and accelerating ovarian ageing, thereby advancing menopause and shortening RL.[2, 24, 49, 50] The nonlinear PIR→ RL association (greater low-to-mid gains, diminishing marginal returns at higher levels) suggests that resource improvements during scarcity-to-sufficiency transitions,such as nutrition, healthcare access and chronic disease management, may yield greater reproductive health benefits. This finding informs targeted interventions but should not supplant causal inference for primary exposures.

Methodologically, we prioritized ANM as the primary outcome to avoid structural coupling with RL.[35] Nonlinearity was assessed using RCS and GAM, with model selection via AIC/joint Wald tests. Exploratory multiple comparisons employed BH-FDR control. Pregnancy loss zero-inflation was addressed via two-part models. Multiple imputation and complex sampling design (weights, stratification, clustering) were rigorously implemented. However, design degrees of freedom and Kish effective sample size may impact power, and that stratification and interaction terms further partition samples, potentially limiting detection of small effects.

Several limitations should be acknowledged, including cross-sectional design and recall bias, which restrict causal inference; possible survivor bias among those >70 years; limited power to detect small effects for pregnancy loss due to measurement and strata sizes; and residual confounding that cannot be completely excluded. Future research should validate temporality and causality in longitudinal cohorts, incorporate objective ovarian reserve markers (e.g., AMH) and multi-omics approaches to elucidate smoking/SES →ovarian aging pathways, and replicate exploratory interactions (e.g., income × pregnancy loss) in larger samples.

## CONCLUSION

Within a nationally representative sample analyzed with complex survey methods and multiple imputation, age at menarche (AAM) showed a robust linear inverse association with reproductive lifespan (RL) (β=−0.9 years per 1-year later AAM) and only a borderline association with age at natural menopause (ANM), indicating that timing of sexual maturation is an important determinant of RL. Parity and pregnancy loss showed no overall associations with ANM or RL. Smoking significantly modified the association between parity and RL, suggesting that lifestyle factors can alter links between reproductive history and reproductive aging. The income-to-poverty ratio exhibited a nonlinear relation with RL, with greater benefits observed in the low-to-middle income range, suggesting that policy interventions could prioritize comprehensive support for populations near the poverty line. Multiple sensitivity analyses and alternative exposure analyses consistently supported the primary findings. However, the lack of statistical significance for some results should be considered in the context of potential power limitations induced by the study design. Future research should aim to validate and extend these findings in longitudinal and mechanistic studies.

## Acknowledgments

The authors thank the NHANES for the openly accessible data.

## Ethics statement

The studies involving humans were approved by the National Center for Health Statistics Ethics Review Board. The studies were conducted in accordance with the local legislation and institutional requirements. The participants provided their written informed consent to participate in this study.

## Author Contributions

JF: Conceptualization, Data curation, Formal analysis, Investigation, Methodology, Visualization, Writing – original draft. JZ: Methodology, Writing – review & editing. JC: Data curation, Formal analysis, Validation. GD: Conceptualization, Funding acquisition, Project administration, Resources, Supervision, Writing – review & editing. All authors contributed to the article and approved the submitted version.

## Data Availability Statement

Publicly available datasets were analyzed in this study. This data can be found here: https://www.cdc.gov/nchs/nhanes

## Figure Legends

**Fig S1. Two-part associations of pregnancy loss with reproductive outcomes.**

Two-part associations of pregnancy loss with reproductive outcomes: left, reproductive lifespan; right, age at menopause. Three adjustment sets (Unadjusted, Basic, Full); shaded bands are 95% CIs. Nonlinearity tests are non-significant and ΔAIC ≈ 1.3–1.4; the linear specification is favored and curves are nearly flat. Analyses use the complex survey design with five imputations pooled via Rubin’s rules.

**Fig S2. Reproductive exposures and outcomes by income strata.**

Rows represent exposures (top: Parity; middle: Gravidity; bottom: Pregnancy loss). Columns represent outcomes (left: Reproductive Lifespan; right: Age at menopause). Survey-weighted, MI-pooled models with the same covariate set as the main analysis. Panel headers include the Wald interaction p-value (exposure × income). Points show Beta and 95% CI.

**Fig S3. Reproductive exposures and outcomes by race strata.**

Rows represent exposures (top: Parity; middle: Gravidity; bottom: Pregnancy loss). Columns represent outcomes (left: Reproductive Lifespan; right: Age at menopause). Survey-weighted, MI-pooled models with the same covariate set as the main analysis. Panel headers include the Wald interaction p-value (exposure × race). Points show Beta and 95% CI.

**Fig S4. Gravidity and reproductive outcomes by smoking status.**

Two-panel forest plot (left: Reproductive Lifespan; right: Age at menopause). Same covariate adjustment as the main analysis. Per-stratum Beta (95% CI) with formatted p-values.

**Fig S5. Pregnancy loss and reproductive outcomes by smoking status.**

Two-panel forest plot (left: Reproductive Lifespan; right: Age at menopause). Same covariate adjustment as the main analysis. Per-stratum Beta (95% CI) with formatted p-values.

